# ETHNICITY, DEPRIVATION, AND CHILDHOOD MORTALITY IN ENGLAND: A COHORT STUDY

**DOI:** 10.1101/2025.03.01.25322994

**Authors:** David Odd, Joanna Garstang, Tom Williams, Sylvia Stoianova, Karen Luyt

**Affiliations:** School of Medicine, Division of Population Medicine, Cardiff University, UK; National Child Mortality Database, Bristol Medical School, University of Bristol, St Michael’s Hospital, Southwell Street, Bristol, UK; School of Nursing and Midwifery, University of Birmingham, Birmingham, UK

**Keywords:** Equity, diversity, inclusion/Health equity, Epidemiology/Epidemiology, Public health/Public health, pediatrics, child health

## Abstract

**OBJECTIVE:** To investigate the relationship between childhood mortality (deaths before 18 years of age) and the ethnicity of the child; and how this is related to local measures of deprivation.

**POPULATION:** All child deaths in England, born at, or over, a gestation of 22 weeks, from April 1, 2019, to March 31, 2023, notified to the NCMD

**DESIGN:** Characteristics and death categories were compared by ethnicity, and the rate of death was calculated using a Poisson distribution. Incident Rate Ratios (IRRs) were derived using White children as a baseline, adjusting for deprivation, age, sex, region of England, and area type.

**MAIN OUTCOME MEASURES:** Rate of death between April 2019 and March 2023.

**RESULTS:** Over the 48 months, a total of 12,142 eligible deaths were reported to the NCMD. After adjusting for confounders, children from Asian (IRR 1.52 (1.45-1.60), Black (IRR 1.61 (1.50-1.73)) and Other ethnicities (IRR 1.12 (1.00-1.25), but not Mixed backgrounds (IRR 0.91 (0.84-1.73)), had a higher rate of death than White children.

**CONCLUSIONS AND RELEVANCE:** In England, children from Asian or Asian British, or Black or Black British backgrounds had the highest rate of death before their 18^th^ birthday, and around 1 in 11 deaths in England would be avoided if all children had the rate of death experienced by White children. However, the relationship between ethnicity and the local environment (esp. deprivation and urban status) was profound, and further work is needed to identify how to best target interventions to reduce the overall inequalities seen.

## INTRODUCTION

Each year in England around 3400 children die before the age of 18 years(1), with recent data suggesting little recent progress(2). Indeed, England has one of the highest infant(3, 4) and child(5) mortality rates in Europe with substantial variation in mortality linked to socioeconomic factors.(6, 7) While ethnicity and race are related but distinct concepts used to categorize people, both the Office for National Statistics (ONS) and the NHS defines ethnicity as a self-identified social group; encompassing a combination of cultural, social, religious, language, and physical factors(8).

While reducing inequity in child mortality should be a national priority, the reasons for child death in high-income countries are complex(9), and further knowledge is needed to implement effective prevention and supportive strategies for those most at risk. The links between social measures of inequality and poor health outcomes are well recognised, but complex.(10, 11) Indeed, while around 65% of the child population are of a White ethnicity, there is substantial variation in the patterns across the regions of England, with additional correlations with local deprivation, making interpretation challenging(12, 13). Recent work has suggested that more than one-fifth of child deaths in England might be avoided if children living in the most deprived areas had the same risk of those living in the least deprived areas(14), with infant mortality rates higher for Black, Asian and mixed ethnicity infants compared to White British infants(15-17). While clear associations with ethnicity are well reported, the patterns and complexities of the relationships are unclear(18, 19), making interventions difficult to design(10, 11). Understanding the reasons for increased child mortality associated with ethnicity and deprivation is key to addressing this long-standing social inequity.

## AIMS

The aim of this work is to investigate the relationship between childhood mortality (deaths before 18 years of age) and the ethnicity of the child; and the patterns and associations which underpin any differences.

## MATERIALS AND METHODS

The National Child Mortality Database (NCMD) collates data on all children who die in England. Data was derived from Child Death Overview Panel (CDOP) death notifications to NCMD. All child deaths were assigned the most likely category of death from the notification data by 3 independent coders (all paediatricians) to identify the most likely category of the cause of death(20). All coders recorded a provisional category of death (see below) or that there was insufficient information provided. For each death, if two or more coders agreed on a category this was taken as the most likely category and where no two coders agreed, the category highest in the following hierarchy was used(21).

1. Suicide
2. Substance Abuse
3. Trauma
4. Malignancy
5. Underlying Medical Condition
6. Intrapartum event
7. Preterm Birth
8. Infection
9. Sudden Unexpected Death in Infancy or Childhood (SUDIC)

In addition, the CDOPs also report baseline characteristics of the child, from which the following data were derived from the notification form:

- Sex of individual (female, male, other (including not known))
- Ethnic Group (Asian or Asian British (Bangladeshi, Chinese, Indian, Pakistani, Any other Asian background), Black or Black British (African, Caribbean, Any other Black background), Mixed (White and Asian, White and Black African, White and Black Caribbean, Any other Mixed background), White or White British (British, Irish, Any other White background (Including Roma, Gypsy or Irish traveller), Other (Arab, Any other ethnic group). Ethnicity is derived from NHS data, and is parent self-reported at source.
- The region of England where the death was reported from
- The gestational age of the child at birth
- From the child’s home postcode at the time of their death, the IMD, a measure of local deprivation(22) (on a score of 1-10) with a lower value suggesting greater deprivation. It is derived at the resolution of the Middle Super Output Area (MSOA) (around 1500 people).

Data was accessed for research purposes on the 7^th^May 2025. Authors had access to information that could identify individual participants data collection.

Please note, that due to an error in the coding of the local deprivation, data was re-analysed on the 9th of March 2026; with resulting attenuated point estimates in the adjusted analysis to the initial pre-print version.

### Statistical Analysis

Deaths of children and young people (CYP) under 18 years of age, occurring from 1^st^April 2019 until 31^st^March 2023 were identified. CYP with a known gestation of less than 22 weeks were excluded. Denominator data (population at risk) were derived from ONS 2021 Census Data(23) through the online portal. A static population at risk was assumed for the study period. Granularity and availability of measures (e.g. age categories) was limited by ONS suppression guidelines. The unadjusted Incident Rate Ratio (IRR) of death compared to White CYP (the largest group) were then derived using a Poisson regression model. Population attributable risks were calculated using CYP of White background as the baseline. (14)Analyses were then repeated, initially adjusting for Index of Multiple Deprivation (IMD) decile and then for all other covariates available (Age (0-4, 5-15 and 16/17 years), Sex (Female or Male), Region (East Midlands, East of England, London, North East, North West, South East, South West, West Midlands, or Yorkshire and the Humber), Area (Rural or Urban)). Selection of age categories was chosen due to available denominator granularity. Next, we repeated the fully adjusted analysis of all deaths, split by the covariates, and tested if the relative rates were different between groups by adding the covariate as an interactive term to the model. The Population Attributable Fractions (PAF) for non-White children vs White children (PAF) were derived from the Poisson models using the Stata ‘punaf’ command.

Finally, as a sensitivity analyses, the main analysis was repeated twice. First, deriving the relative rates for each ethnic group, compared to CYP from White backgrounds, split by each year of age, and adjusting for the above covariates (Region, Deprivation Decile and Area) except sex (due to limitations in available data). Secondly, the analysis was repeated, this time deriving relative rates for all deaths, and categories of death, split by a more granular measure of ethnicity.

Data are presented as number (%), rate per 100,000 person years (95% confidence interval (CI)), incident rate ratio (IRR) (95% CI), or PAF (95% CI). Analysis was performed using Stata version 17. All models were tested to support the use of a Poisson distribution. Cell counts less than 5 were suppressed to maintain anonymity of potentially identifiable data.

## RESULTS

Over the 48 months, a total of 13,743 deaths were reported to NCMD, or which 915 were excluded for birth before 22 weeks gestation, leaving 12,828 deaths. A total of 686 were removed due to missing ethnicity data, leaving 12,142 for the main analysis.

The highest number of deaths was seen in White CYP (n=7758), followed by Asian (n=2242), Black (n=1039), Mixed (n=742) and Other (n=361) (eTable 1). The suspected cause of death also appeared to vary by ethnicity (p<0.001) (eTable 2), although Underlying Disease (3840/11872 (32.3%)) and prematurity (2676/11872 (22.5%)) were the highest two groups for all ethnicities.

Rate of death varied by ethnicity (p<0.001) (Table 1). The lowest rate of death was seen by CYP of a white background (22.71 (22.20-23.22)), while the highest rate was seen by Black or Black British (38.68 (36.36-41.10)), and Asian or Asian British (38.66 (37.09-40.30)) CYP. The rate of all-cause mortality was different by ethnicity for deaths by malignancy (p=0.0092), preterm birth (p<0.001), infection (p<0.001), trauma (p<0.001), substance abuse (p=0.0034), suicide (p=0.0061), SUDIC (p=0.0001) and underlying disease (p<0.001).

**Table 1.**
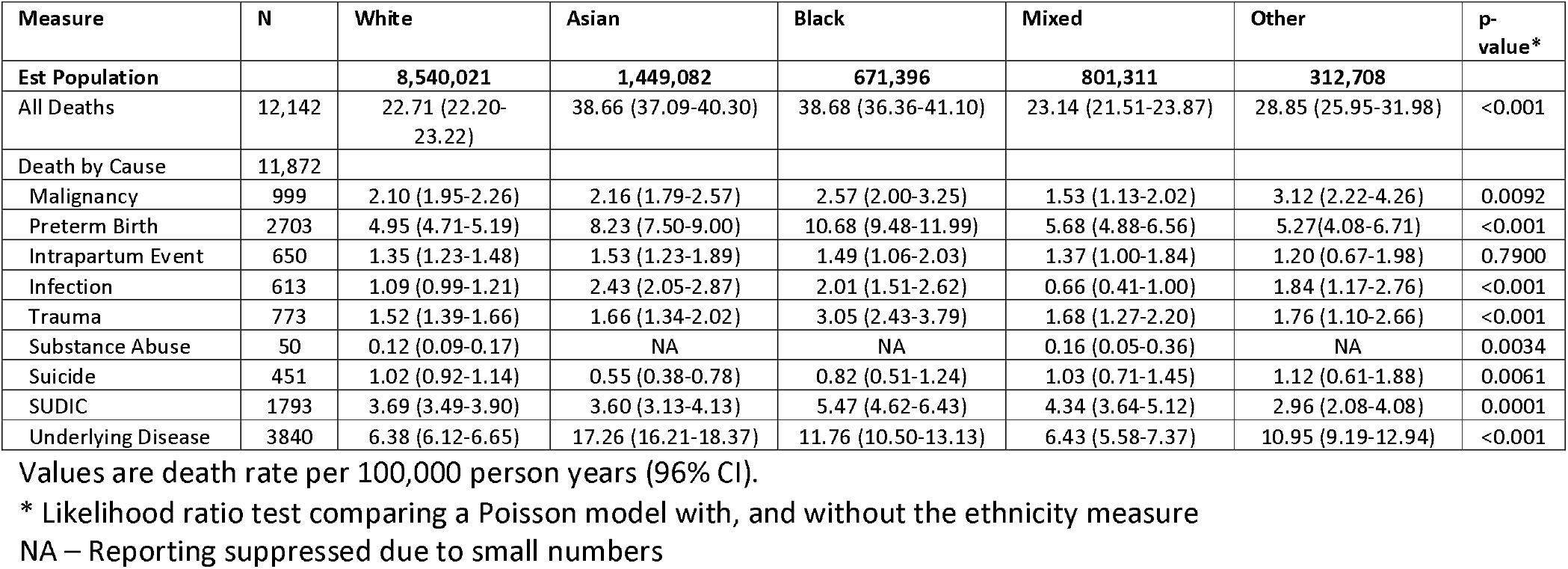
Rate of death (per 100,000 CYP per year) for CYP in England between April 2019 and March 2023; split by ethnicity and provisional cause.

| Measure | N | White | Asian | Black | Mixed | Other | p-value * |
| --- | --- | --- | --- | --- | --- | --- | --- |
| **Est Population** |  | **8,540,021** | **1,449,082** | **671,396** | **801,311** | **312,708** |  |
| All Deaths | 12,142 | 22.71 (22.20-23.22) | 38.66 (37.09-40.30) | 38.68 (36.36-41.10) | 23.14 (21.51-23.87) | 28.85 (25.95-31.98) | <0.001 |
| Death by Cause | 11,872 |  |  |  |  |  |  |
| Malignancy | 999 | 2.10 (1.95-2.26) | 2.16 (1.79-2.57) | 2.57 (2.00-3.25) | 1.53 (1.13-2.02) | 3.12 (2.22-4.26) | 0.0092 |
| Preterm Birth | 2703 | 4.95 (4.71-5.19) | 8.23 (7.50-9.00) | 10.68 (9.48-11.99) | 5.68 (4.88-6.56) | 5.27 (4.08-6.71) | <0.001 |
| Intrapartum Event | 650 | 1.35 (1.23-1.48) | 1.53 (1.23-1.89) | 1.49 (1.06-2.03) | 1.37 (1.00-1.84) | 1.20 (0.67-1.98) | 0.7900 |
| Infection | 613 | 1.09 (0.99-1.21) | 2.43 (2.05-2.87) | 2.01 (1.51-2.62) | 0.66 (0.41-1.00) | 1.84 (1.17-2.76) | <0.001 |
| Trauma | 773 | 1.52 (1.39-1.66) | 1.66 (1.34-2.02) | 3.05 (2.43-3.79) | 1.68 (1.27-2.20) | 1.76 (1.10-2.66) | <0.001 |
| Substance Abuse | 50 | 0.12 (0.09-0.17) | NA | NA | 0.16 (0.05-0.36) | NA | 0.0034 |
| Suicide | 451 | 1.02 (0.92-1.14) | 0.55 (0.38-0.78) | 0.82 (0.51-1.24) | 1.03 (0.71-1.45) | 1.12 (0.61-1.88) | 0.0061 |
| SUDIC | 1793 | 3.69 (3.49-3.90) | 3.60 (3.13-4.13) | 5.47 (4.62-6.43) | 4.34 (3.64-5.12) | 2.96 (2.08-4.08) | 0.0001 |
| Underlying Disease | 3840 | 6.38 (6.12-6.65) | 17.26 (16.21-18.37) | 11.76 (10.50-13.13) | 6.43 (5.58-7.37) | 10.95 (9.19-12.94) | <0.001 |

Values are death rate per 100,000 person years (96% CI).
\* Likelihood ratio test comparing a Poisson model with, and without the ethnicity measure
NA – Reporting suppressed due to small numbers

**Table 2.** Incident rate ratios (IRR) of CYP death by ethnicity, stratified by provisional cause.

| Measure | n | Incident Rate Ratio (IRR) (vs White ethnicity) |  |  |  |  | PAF |
| --- | --- | --- | --- | --- | --- | --- | --- |
|  |  | White | Asian | Black | Mixed | Other |  |
| **Estimated Population at Risk** |  | **8,540,021** | **1,449,082** | **671,396** | **801,311** | **312,708** |  |
| **Unadjusted IRR** |  |  |  |  |  |  |  |
| All Deaths | 12,142 | 1 (ref) | 1.70 (1.62-1.78) | 1.70 (1.60-1.82) | 1.02 (0.95-1.10) | 1.27 (1.14-1.41) | 11.9% (10.7% to 13.1%) |
| Malignancy | 999 | 1 (ref) | 1.03 (0.85-1.24) | 1.22 (0.96-1.57) | 0.73 (0.55-0.97) | 1.49 (1.08-2.05) | 1.0% (-2.9% to 4.8%) |
| Preterm Birth | 2703 | 1 (ref) | 1.66 (1.50-1.84) | 2.16 (1.90-2.45) | 1.15 (0.98-1.34) | 1.07 (0.83-1.36) | 13.7% (11.2% to 16.2%) |
| Intrapartum Event | 650 | 1 (ref) | 1.14 (0.90-1.42) | 1.10 (0.80-1.52) | 1.02 (0.75-1.38) | 0.89 (0.53-1.48) | 2.0% (-2.9% to 6.7%) |
| Infection | 613 | 1 (ref) | 2.22 (1.83-2.70) | 1.84 (1.38-2.44) | 0.60 (0.39-0.93) | 1.68 (1.10-2.56) | 15.9% (10.4% to 20.4%) |
| Trauma | 773 | 1 (ref) | 1.09 (0.88-1.36) | 2.01 (1.592-54) | 1.11 (0.84-1.47) | 1.16 (0.76-1.77) | 7.4% (2.7% to 11.9%) |
| Substance Abuse | 50 | 1 (ref) | NA | NA | NA | NA | NA |
| Suicide | 451 | 1 (ref) | 0.54 (0.38-0.77) | 0.80 (0.52-1.23) | 1.00 (0.70-1.44) | 1.09 (0.64-1.86) | -7.0% (-12.4% to 1.8%) |
| SUDIC | 1793 | 1 (ref) | 0.98 (0.84-1.13) | 1.48 (1.25-1.76) | 1.17 (0.99-1.40) | 0.80 (0.58-1.11) | 3.0% (0.7% to 5.9%) |
| Underlying Disease | 3840 | 1 (ref) | 2.71 (2.51-2.92) | 1.84 (1.64-2.07) | 1.01 (0.87-1.16) | 1.72 (1.44-2.04) | 21.7% (19.5% to 23.9%) |
| **Adjusted for Deprivation** |  |  |  |  |  |  |  |
| All Deaths | 12,142 | 1 (ref) | 1.48 (1.41-1.55) | 1.42 (1.33-1.51) | 0.97 (0.90-1.04) | 1.05 (0.95-1.18) | 8.3% (7.0% to 9.3%) |
| Malignancy | 999 | 1 (ref) | 1.04 (0.85-1.26) | 1.22 (0.95-1.58) | 0.73 (0.55-0.98) | 1.27 (0.89-1.80) | 0.4% (-3.6% to 4.3%) |
| Preterm Birth | 2703 | 1 (ref) | 1.43 (1.29-1.59) | 1.78 (1.56-2.02) | 1.09 (0.93-1.26) | 0.92 (0.72-1.18) | 10.1% (7.3% to 12.7%) |
| Intrapartum Event | 650 | 1 (ref) | 1.10 (0.87-1.39) | 1.05 (0.75-1.46) | 1.00 (0.73-1.37) | 0.85 (0.51-1.43) | 1.1% (-4.0% to 6.0%) |
| Infection | 613 | 1 (ref) | 1.99 (1.63-2.44) | 1.59 (1.18-2.13) | 0.57 (0.37-0.89) | 1.44 (0.93-2.22) | 12.9% (7.1% to 18.4%) |
| Trauma | 773 | 1 (ref) | 0.92 (0.74-1.15) | 1.62 (1.27-2.05) | 1.04 (0.79-1.38) | 0.85 (0.54-1.35) | 2.8% (-2.3% to 7.6%) |
| Substance Abuse | 50 | 1 (ref) | NA | NA | NA | NA | NA |
| Suicide | 451 | 1 (ref) | 0.55 (0.38-0.79) | 0.82 (0.53-1.27) | 1.02 (0.71-1.45) | 1.11 (0.65-1.90) | -6.3% (-12.0% to -0.1%) |
| SUDIC | 1793 | 1 (ref) | 0.75 (0.65-0.88) | 1.08 (0.90-1.27) | 1.07 (0.90-1.27) | 0.63 (0.45-0.87) | -3.9% (-7.2% to -0.7%) |
| Underlying Disease | 3840 | 1 (ref) | 2.35 (2.17-2.54) | 1.52 (1.35-1.72) | 0.96 (0.83-1.10) | 1.41 (1.18-1.69) | 18.4% (16.0% to 20.7%) |
| **Fully Adjusted\*** |  |  |  |  |  |  |  |
| All Deaths | 12,142 | 1 (ref) | 1.52 (1.45-1.60) | 1.61 (1.50-1.73) | 0.91 (0.84-1.73) | 1.12 (1.00-1.25) | 8.9% (7.6% to 10.2%) |
| Malignancy | 999 | 1 (ref) | 1.04 (0.85-1.27) | 1.23 (0.95-1.61) | 0.73 (0.55-0.98) | 1.27 (0.89-1.81) | 0.3% (-4.0% to 4.4%) |
| Preterm Birth | 2703 | 1 (ref) | 1.37 (1.23-1.52) | 1.77 (1.55-2.03) | 1.06 (0.91-1.24) | 0.91 (0.71-1.16) | 9.1% (6.2% to 11.9%) |
| Intrapartum Event | 650 | 1 (ref) | 1.02 (0.80-1.29) | 0.93 (0.66-1.30) | 0.94 (0.68-1.28) | 0.77 (0.46-1.30) | -1.4% (-7.0% to 4.0%) |
| Infection | 613 | 1 (ref) | 2.10 (1.70-2.58) | 1.76 (1.30-2.40) | 0.55 (0.36-0.86) | 1.54 (0.99-2.39) | 13.8% (7.8% to 19.3%) |
| Trauma | 773 | 1 (ref) | 0.96 (0.76-1.20) | 1.69 (1.31-2.18) | 1.11 (0.83-1.47) | 1.11 (0.83-1.47) | 3.8% (-1.6% to 8.8%) |
| Substance Abuse | 50 | 1 (ref) | NA | NA | NA | NA | NA |
| Suicide | 451 | 1 (ref) | 0.59 (0.40-0.85) | 0.92 (0.59-1.45) | 0.98 (0.67-1.42) | 1.22 (0.71-2.11) | -5.1% (-11.1% to 0.5%) |
| SUDIC | 1793 | 1 (ref) | 0.77 (0.66-0.89) | 1.20 (1.00-1.44) | 0.98 (0.82-1.17) | 0.65 (0.47-0.90) | -3.6% (-7.1% to -0.2%) |
| Underlying Disease | 3840 | 1 (ref) | 2.49 (2.29-2.70) | 1.81 (1.59-2.05) | 0.91 (0.78-1.05) | 1.55 (1.30-1.86) | 19.7% (17.3% to 22.0%) |

Values are incident rate ratios (IRR) (95% CI) or population attributable fraction (95% CI)
\* Adjusted for local deprivation, age at death, sex, region of England and area (Rural or Urban).
NA – Reporting suppressed due to small numbers

In the fully adjusted analysis, the association of mixed ethnicity weakened slightly (IRR 0.91 (0.84-1.73)), but remained for all other ethnic groups, compared to White CYP. Children of Asian backgrounds had lower rate of dying by suicide (IRR 0.59 (0.40-0.85)) and SUDIC (0.77 (0.66-0.89)), while those of mixed backgrounds had a lower rate of infection (IRR 0.55 (0.36-0.86)) and malignancy (IRR 0.73 (0.55-0.98)). In the adjusted model the PAF, for non-White vs White children, was 8.9% (7.6% to 10.2%); around 1 in 11 deaths. A raised PAF was also seen for death after preterm birth, by infection, and underlying disease. SUDIC had a negative PAF (compared to white children) of −3.6% (−7.1% to −0.2%).

Differences in rates of death across ethnicities varied by age (p<0.001), region (p<0.001), area (p=0.015) and deprivation (p<0.001); but not sex (p=0.113) (Table 3) (Figure 1). The associations for all ethnic groups appeared to attenuate as the age group increased, with no associations seen in the oldest age range. Patterns of mortality appeared to vary by area, with less associations seen for children in rural areas. Rate of death in different ethnic groups also varied by region and deprivation, although patterns were complex; the highest relative rate was seen in Black and Asian CYP in deciles 5/6 (i.e. the middle 2 deciles). For regional changes, the highest relative rate for Asian CYP were seen in Yorkshire and Humber (IRR 1.87 (1.63-2.15)), and the North East (IRR 1.86 (1.41-2.26)), while relative rate was lowest in the South West (IRR 0.95 (0.66-1.36)). Highest relative rate for Black CYP were seen in London (IRR 1.75-2.23), and the South East (IRR 1.91 (1.56-2.35). The relative rate was lowest, compared to White children, in the North West (IRR 1.07 (0.85-1.25)). The number, and hence precision, made interpreting the relative rates for children from mixed or other background difficult, although children from mixed backgrounds appeared to have a lower risk of death in the North West (IRR 0.72 (0.57-0.92)), while those from Other backgrounds, a higher risk in the South West (IRR 1.67 (1.04-2.66)). When repeating the analysis, split by year of age, relative rates for Asian and Asian British CYP, and for Black and Black British CYP were mostly higher than white CYP until age 7 (with one similar IRR at age 3 for Black CYP (IRR 0.80 (0.42-1.52)) (eTable 3) (eFigure 1). Children from mixed or other backgrounds had variable relative rates across the age range.

**Table 3.** Incident Rate Ratio (IRR) of CYP death by ethnicity, stratified by CYP characteristics.

| Measure | N | White | Asian | Black | Mixed | Other | P<sub>interaction</sub> * |
| --- | --- | --- | --- | --- | --- | --- | --- |
| Est Population |  | 8,540,021 | 1,449,082 | 671,396 | 801,311 | 312,708 |  |
| Age | 12,142 |  |  |  |  |  | <0.001 |
| 0-4 years | 8569 | 1 (Ref) | 1.57 (1.47-1.66) | 1.72 (1.59-1.87) | 0.90 (0.82-1.98) | 1.11 (0.97-1.26) |  |
| 5-15 years | 2537 | 1 (Ref) | 1.58 (1.41-1.76) | 1.47 (1.25-1.72) | 0.94 (0.79-1.12) | 1.29 (1.03-1.62) |  |
| 16-17 years | 1036 | 1 (Ref) | 1.08 (0.89-1.31) | 1.17 (0.91-1.51) | 0.92 (0.69-1.23) | 0.82 (0.54-1.26) |  |
| Sex | 12,108 |  |  |  |  |  | 0.113 |
| Female | 5210 | 1 (Ref) | 1.66 (1.54-1.79) | 1.66 (1.50-1.85) | 0.90 (0.80-1.02) | 1.17 (0.99-1.38) |  |
| Male | 6898 | 1 (Ref) | 1.42 (1.33-1.52) | 1.57 (1.43-1.72) | 0.91 (0.82-1.00) | 1.08 (0.94-1.25) |  |
| Region | 12,142 |  |  |  |  |  | <0.001 |
| East Midlands | 1012 | 1 (Ref) | 1.26 (1.04-1.52) | 1.61 (1.24-2.10) | 1.17 (0.92-1.49) | 0.68 (0.39-1.18) |  |
| East of England | 1165 | 1 (Ref) | 1.48 (1.23-1.77) | 1.80 (1.42-2.28) | 0.93 (0.73-1.17) | 1.34 (0.91-1.99) |  |
| London | 2006 | 1 (Ref) | 1.51 (1.35-1.69) | 1.97 (1.75-2.23) | 0.86 (0.73-1.02) | 1.19 (0.98-1.45) |  |
| North East | 602 | 1 (Ref) | 1.86 (1.41-2.26) | 1.54 (0.97-2.45) | 0.43 (0.22-0.87) | 1.58 (0.96-2.61) |  |
| North West | 1762 | 1 (Ref) | 1.42 (1.23-1.61) | 1.07 (0.85-1.35) | 0.72 (0.57-0.92) | 0.95 (0.70-1.31) |  |
| South East | 1640 | 1 (Ref) | 1.61 (1.39-1.86) | 1.91 (1.56-2.35) | 0.95 (0.78-1.16) | 1.18 (0.83-1.69) |  |
| South West | 914 | 1 (Ref) | 0.95 (0.66-1.36) | 1.57 (1.09-2.27) | 0.86 (0.63-1.18) | 1.67 (1.04-2.66) |  |
| West Midlands | 1652 | 1 (Ref) | 1.45 (1.28-1.64) | 1.32 (1.09-1.64) | 0.98 (0.81-1.19) | 1.16 (0.88-1.53) |  |
| Yorkshire and the Humber | 1329 | 1 (Ref) | 1.87 (1.63-2.15) | 1.66 (1.29-2.15) | 0.94 (0.72-1.22) | 0.78 (0.51-1.20) |  |
| Area | 12,085 |  |  |  |  |  | 0.0149 |
| Urban | 10,705 | 1 (Ref) | 1.55 (1.48-1.63) | 1.65 (1.54-1.77) | 0.92 (0.85-1.00) | 1.14 (1.02-1.27) |  |
| Rural | 1380 | 1 (Ref) | 0.84 (0.55-1.30) | 1.17 (0.69-1.98) | 0.77 (0.57-1.04) | 0.95 (0.45-1.99) |  |
| Deprivation | 12,081 |  |  |  |  |  | <0.001 |
| 1/2 (Most Deprived) | 4132 | 1 (Ref) | 1.17 (1.08-1.26) | 1.26 (1.14-1.40) | 0.84 (0.74-0.96) | 0.80 (0.67-0.95) |  |
| 3/4 | 2749 | 1 (Ref) | 1.98 (1.79-2.19) | 2.17 (1.90-2.47) | 0.97 (0.83-1.14) | 1.43 (1.15-1.76) |  |
| 5/6 | 2110 | 1 (Ref) | 2.13 (1.88-2.41) | 2.36 (1.99-2.80) | 0.88 (0.72-1.06) | 1.22 (0.92-1.62) |  |
| 7/8 | 1679 | 1 (Ref) | 1.49 (1.27-1.76) | 2.13 (1.68-2.70) | 0.98 (0.80-1.20) | 1.47 (1.06-2.05) |  |
| 9/10 (Least Deprived) | 1411 | 1 (Ref) | 1.41 (1.17-1.69) | 1.01 (0.66-1.54) | 0.92 (0.74-1.14) | 1.79 (1.26-2.53) |  |

Values are IRR (95% CI)
Adjusted for deprivation decile (IMD), age at death, sex, region of England and area (Rural or Urban).
\* p-value is derived by the Likelihood ratio test, testing the model with, or without an interaction term for the covariate.

**Figure 1.**
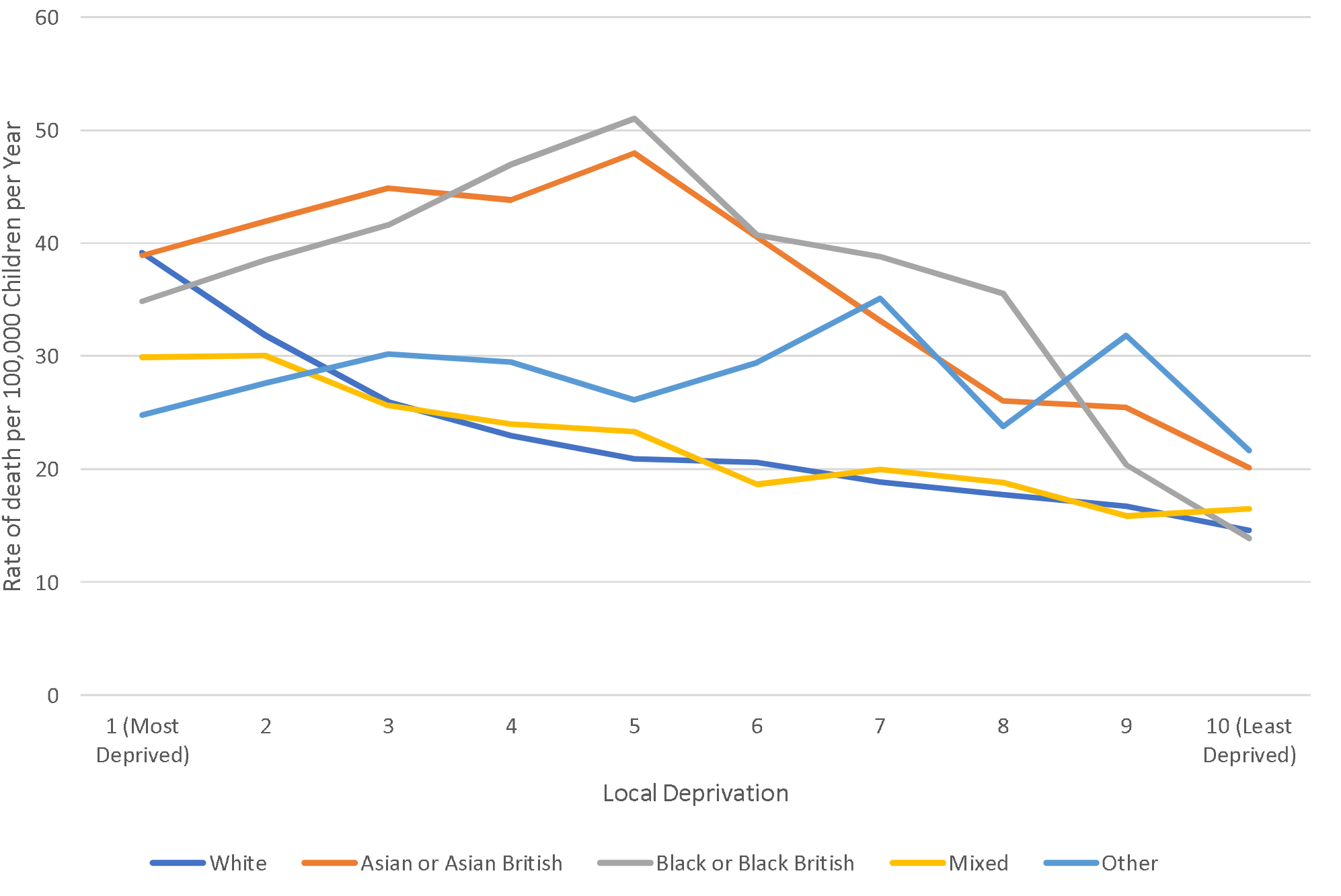
Unadjusted rate of death per 100,000 person years, by ethnic group, and local deprivation. Deprivation measured using the Index of Multiple Deprivation (Office of National Statistics)

A further 20 (0.2%) of children had missing data on the more granular measure of ethnicity (Table 4&eTable 4), making a total analysis cohort of 12,122 (94.5%) children. In addition, absolute numbers became small, making identifiability of data and interpretation problematic. Compared to White British children, children from Pakistani (IRR 2.02 (1.89-2.16)), other Asian (IRR 2.05 (1.86-2.27) and Irish CYP (IRR 2.56 (1.99-3.30)) backgrounds had the highest relative rates of death, followed by those from African (IRR 1.86 (1.72-2.02))) and Caribbean communities (IRR 1.87 (1.59-2.19)). Children from Gypsy or Irish traveller (IRR 1.45 (1.02-2.08)), White-Other (IRR 1.49 (1.39-1.61), Bangladeshi (RR 1.28 (1.13-1.46)), Black-Other (IRR 1.22 (1.04-1.44)), Mixed Other (IRR 1.21 (1.06-1.38)), and Other (IRR 1.46 (1.30-1.65)), also appeared to have rates higher than White British children. Children from Chinese (IRR 0.73 (0.53-0.99), Mixed (White-Asian) (IRR 0.77 (0.67-0.90)), and Arab (IRR 0.56 (0.42-0.73)), had lower rates than White-British children.

**Table 4.**
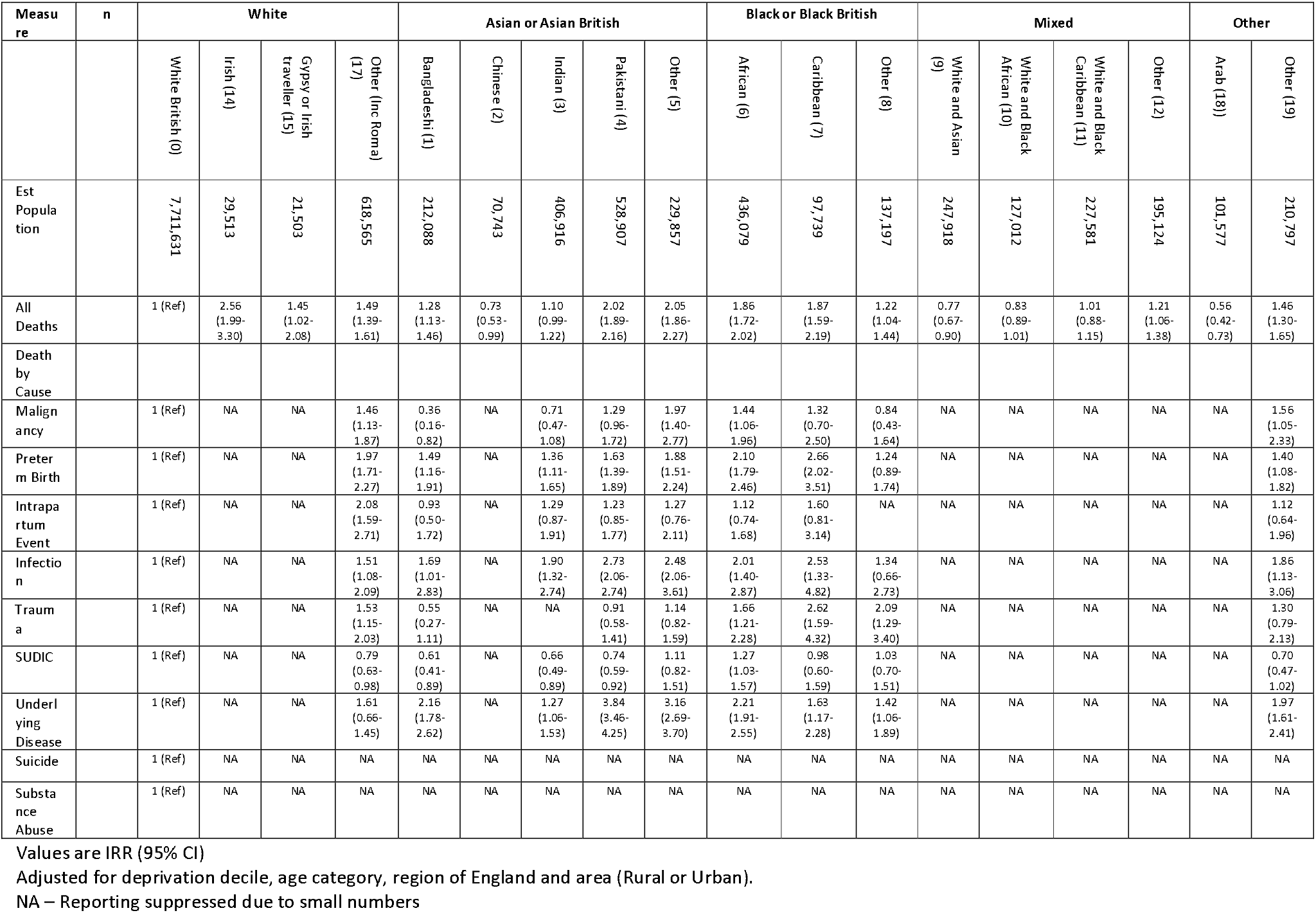
Incident Rate Ratio (IRR) for death of CYP in England between April 2019 and March 2023; split by ethnicity (n=12,122)

## DISCUSSION

### Key Findings

The patterns of mortality in England by ethnicity are complex. White children have much lower risks than those of Black or Asian children, with mortality 50% lower, not explained by measures of local deprivation or demographics. However, the relative components across categories and different ethnic groups was complex. Children from Bangladeshi and Pakistani backgrounds had relatively high risks of death from underlying diseases, a category predominantly driven by genetic disease and congenital abnormalities(14, 24), while Caribbean and Black-Other children had 2 to 3-fold increased rate of death from trauma, preterm birth and infection (compared to White-British children). The patterns of risk also appeared to vary with age, with the highest difference seen in the youngest. The relationship between mortality and ethnicity also appeared strongest in urban areas, and the lack of a clear impact for families living in more affluent areas, supports a substantial environmental, and modifiable, element to these inequalities.

### In Context

Overall, this work presents similar findings of higher child mortality for minoritised populations in other countries, with adverse environments having a direct impact on child mortality(17, 25), and expands on previous work looking at infant mortality(14). The biggest contributors to inequalities appeared to be death after preterm birth, death from infections, and underlying disease. However, the patterns seen are complex, with rates varying within, and between, groups. While we reported higher risks for Black or Black British children for most areas, the profile varied across categories, with some (e.g. suicide) similar to the population average, while others substantially higher. In particular, children from mixed and Asian backgrounds demonstrated a substantial variation within their wider classification. Children from the Pakistani, Indian and Chinese communities had widely different mortality, with the later having the lowest risk of death in this work. Within the Mixed category, children of White and Asian backgrounds, had a lower risk of death than White-British children, while those of White and Caribbean (or Other), having raised risks. Children of White, but not White-British backgrounds, had relative rates similar to those seen in Asian infants for many categories of death. Equally, the attenuation of ethnic patterning in rural areas was striking; suggesting that any impacts or pathways may be different in areas where non-White groups are less prevalent. Finally, the relationship with deprivation, was complicated, although in the least deprived areas, two groups (Asian and Other) continued to have higher risks of death than White children.

One commonly raised cause of death, and a large contributor to overall mortality is that of preterm birth(14, 26), with broad impacts on childhood outcomes(27), and improving maternal health may be the key to reducing broader childhood inequalities. The patterns of infectious disease deaths are consistent with patterns seen in the COVID-19 pandemic(15), and may reflect differences in the risks of acquiring the infection or access to healthcare for therapeutics(28). Finally, deaths from underlying disease in this population is predominantly driven by genetic disorders or congenital abnormalities, and broad population interventions, would be needed to reduce the incidence and impact. Further work is underway to further understand this group. Underpinning the differences across ages, is likely the relative impacts of different pathologies, with Suicide and Trauma having a greater impact in the older children, and very different patterns with ethnicity.

A recent systematic review identified contextual (e.g. deprivation) and individual (e.g. ethnicity) characteristics affecting material circumstances, behaviour, psychosocial environment, and healthcare utilisation, and leading to adverse birth outcomes(29). In contrast to the worsening mortality in the UK, in the USA, the gap between outcomes for Black, Hispanic and White infants is decreasing(30), although infant mortality of black college-educated parents remains higher than White infants(31). Access to healthcare is a major issue(32) while refugee children may be lost to medical services(33), with barriers experienced through language, health literacy and transport inequalities(34), and disjointed healthcare more likely delivered through Emergency Departments(35). Mothers from ethnic minorities report negative experiences related to poor communication, cultural insensitivity and disregard of their needs(36), and direct racism(37); while cultural competence training programmes may have limited benefits(38). However, the relationship between ethnicity and the local environment, in particular the measures of deprivation and urban status were profound, with children from least deprived areas having little to no patterning of mortality by their ethnicity. A similar effect was seen for children in rural areas; where no association with any ethnic group was identified.

### Limitations

This work is based on the ongoing data collection from the NCMD. While we had some missing data on ethnicity, data completeness was generally good, and the primary analysis was based on the statutory reporting of deaths. This missing, or misallocation of ethnicity is likely the biggest limitation of this work, further complicated by incomplete national data. Whether the missing data in the denominator introduces, or minimises, bias in this work in unknown, but the possible discordance between data sources is likely to have some impact on the point estimates seen here(39). We also were unable to account for clustering within families, and so some over-correction in the adjusted analyses is possible. We have used ONS data to derive the population at risk, and this was estimated at a mid-point of the study. While changes in population number and structure may have biased our estimates it is unlikely that they could have accounted for the associations ween here, although absolute rates should be interpreted with caution. Fortunately, child death in England is rare, particularly for causes such as suicide and substance abuse, which may further limit interpretation; particularly in the stratified analyses. Finally, due to the statutory nature of the CDOP process we anticipate being aware of all deaths of children normally resident in England, with previous validation work supporting this(2). However results from each category of death in this work is provisional on data available to the team at the notification, and may change as further information is provided during the subsequent CDOP investigations and report.

## Conclusions

In England, children from Asian or Asian British, or Black or Black British backgrounds had the highest rate of death before their 18^th^ birthday, and around 1 in 11 deaths in England would be avoided if all children had the rate of death experienced by White children. This association was highest in the youngest children and in urban areas. Causes of this increased mortality included Underlying Disease, Infection and Preterm Birth, and unexplained variation in inequalities was seen across England. However, the relationship between ethnicity and the local environment (e.g. deprivation and urban status) was profound, and further work is needed to clarify this intersectionality, and identify how to best target interventions to reduce the overall inequalities seen.

### Patient and Public Involvement

Parent and public involvement guided the design and setting up of the NCMD at establishment and real-time child mortality surveillance system at the beginning of the COVID-19 pandemic.

## Supporting information

eFigure 1

eTable 1

eTable 2

eTable 3

eTable 4

## Acknowledgements

We thank all Child Death Overview Panels (CDOPs) who submitted data for the purposes of this report and all child death review professionals for submitting data and providing additional information when requested.

Parent and public involvement is at the heart of the NCMD programme. We are indebted to Charlotte Bevan (Sands - Stillbirth and Neonatal Death Charity), Ann Chalmers (Child Bereavement UK) and Jenny Ward (Lullaby Trust), who represent bereaved families on the NCMD programme steering group.

We thank the NCMD team for technical and administrative support.

David Odd had full access to all the data in the study and takes responsibility for the integrity of the data and the accuracy of the data analysis.

## Ethics approval and consent to participate

The NCMD legal basis to collect confidential and personal level data under the Common Law Duty of Confidentiality has been established through the Children Act 2004 Sections M - N, Working Together to Safeguard Children 2018 (https://consult.education.gov.uk/child-protection-safeguarding-and-family-law/working-together-to-safeguard-children-revisions-t/supporting_documents/Working%20Together%20to%20Safeguard%20Children.pdf) and associated Child Death Review Statutory &Operational Guidance https://assets.publishing.service.gov.uk/government/uploads/system/uploads/attachment_data/file/859302/child-death-review-statutory-and-operational-guidance-england.pdf).

The NCMD legal basis to collect personal data under the General Data Protection Regulation (GDPR) without consent is defined by GDPR Article 6 (e) Public task and 9 (h) Health or social care (with a basis in law).

## Authors Contributions

DO: I declare that I contributed to conceptualization, data curation, formal analysis, funding acquisition, investigation, methodology, writing the original draft and review and editing of the final version.

JG: I declare that I contributed to conceptualization, data curation, funding acquisition, investigation, methodology, writing the original draft and review and editing of the final version.

SS: I declare that I contributed to conceptualization, data curation, funding acquisition, investigation, methodology, writing the original draft and review and editing of the final version.

TW: I declare that I contributed to conceptualization, data curation, funding acquisition, investigation, methodology, writing the original draft and review and editing of the final version.

KL: I declare that I contributed to conceptualization, data curation, funding acquisition, investigation, methodology, writing the original draft and review and editing of the final version.

## Transparency statement

Prof Karen Luyt (the manuscript’s guarantor) affirms that the manuscript is an honest, accurate, and transparent account of the study being reported; that no important aspects of the study have been omitted; and that any discrepancies from the study as originally planned have been explained.

## Availability of data

Aggregate data may be available on request to the corresponding author, and subject to approval by HQIP.

## REFERENCES

1. OddD, StoianovaS, WilliamsT, FlemingP, LuytK. Child mortality in England during the first year of the COVID-19 pandemic. Arch Dis Child. 2022;107(3):e22.

2. OddD, StoianovaS, WilliamsT, FlemingP, LuytK. Child mortality in England after national lockdowns for COVID-19: An analysis of childhood deaths, 2019-2023. PLoS Med. 2025;22(1):e1004417.

3. MacDormanMF, MatthewsTJ, MohangooAD, ZeitlinJ. International comparisons of infant mortality and related factors: United States and Europe, 2010. National vital statistics reports : from the Centers for Disease Control and Prevention, National Center for Health Statistics, National Vital Statistics System. 2014;63(5):1–6.

4. LozanoR, FullmanN, MumfordJE, KnightM, BarthelemyCM, AbbafatiC, et al.Measuring universal health coverage based on an index of effective coverage of health services in 204 countries and territories, 1990–2019: a systematic analysis for the Global Burden of Disease Study 2019. The Lancet. 2020;396(10258):1250–84.

5. Statistics OfN. UK drops in European child mortality rankings. 2017.

6. ZylbersztejnA, GilbertR, HjernA, WijlaarsL, HardelidP.Child mortality in England compared with Sweden: a birth cohort study. Lancet(London, England). 2018;391(10134):2008–18.

7. OddD, StoianovaS, WilliamsT, OddD, KurinczukJJ, WolfeI, et al.What is the relationship between deprivation, modifiable factors and childhood deaths: a cohort study using the English National Child MortalityDatabase. BMJ Open. 2022;12(12):e066214.

8. Ethnicity and National Identity in England and Wales: 2011. Office for National Statistics (UK). 2012.

9. SidebothamP, FraserJ, CovingtonT, FreemantleJ, PetrouS, Pulikottil-JacobR, et al.Understanding why children die in high-income countries. Lancet. 2014;384(9946):915–27.

10. OddD, WIlliamsT, StoianovaS, SleapV, GloverN, RossouwG, et al.The Contribution of Newborn Health to Child Mortality across England.: National Child Mortality Database (UK); 2022[Available from: <https://www.ncmd.info/wp-content/uploads/2022/07/Perinatal-FINAL.pdf. >

11. ZangE, ZhengH, YangYC, LandKC. Recent trends in US mortality in early and middle adulthood: racial/ethnic disparities in inter-cohort patterns. International journal of epidemiology. 2019;48(3):934–44.

12. OddD, StoianovaS, WilliamsT, OddD, KurinczukJJ, WolfeI, et al.What is the relationship between deprivation, modifiable factors and childhood deaths: a cohort study using the English National Child Mortality Database. BMJ Open. 2022;12(12):e066214.

13. OddD, StoianovaS, SleapV, WilliamsT, CookN, McGeehanL, et al.Child Mortality and Social Deprivation. National Child Mortality Database (UK). 2021.

14. OddDE, StoianovaS, WilliamsT, OddD, Edi-OsagieN, McClymontC, et al.Race and Ethnicity, Deprivation, and Infant Mortality in England, 2019-2022. JAMA Netw Open. 2024;7(2):e2355403.

15. OddD, StoianovaS, WilliamsT, FlemingP, LuytK. Child Mortality in England after the Pandemic. Increasing Mortality and Inequalities. medRxiv. 2024:2024.05.24.24307855.

16. FloresG, EscalaMK, HallBG. Dead wrong: the growing list of racial/ethnic disparities in childhood mortality. J Pediatr. 2015;166(4):790–3.

17. PedersenGS, MortensenLH, AndersenAM. Ethnic variations in mortality in pre-school children in Denmark, 1973-2004. Eur J Epidemiol. 2011;26(7):527–36.

18. Ethnic Inequalities in Healthcare: A Rapid Evidence Review. NHS Race and Health Observatory; 20222022.

19. Mapping of Existing Policy Interventions to Tackle Ethnic Health Inequalities in Maternity and Neonatal Health in England: A Systematic Scoping Review with Stakeholder Engagement. NHS Race & Health Observatory; 2022.

20. OddD, StoianovaS, SleapV, WilliamsT, CookN, McGeehanL, et al.Child Mortality and Social Deprivation. 2021.

21. Child Death Review: Statutory and Operational Guidance (England). HM Government (UK). London; 2018.

22. English indices of deprivation 2019: technical report. Ministry of Housing, Communities & Local Government (UK); 2019.

23. Create a custom dataset (Census 2021): Office for National Statistics (ONS); 2022[Available from: <https://www.ons.gov.uk/datasets/create. >

24. OddD, StoianovaS, WilliamsT, FlemingP, LuytK. Child Mortality in England During the First 2 Years of the COVID-19 Pandemic. JAMA Netw Open. 2023;6(1):e2249191.

25. WolfER, RivaraFP, OrrCJ, SenA, ChapmanDA, WoolfSH. Racial and Ethnic Disparities in All-Cause and Cause-Specific Mortality Among US Youth. Jama. 2024;331(20):1732–40.

26. UK Perinatal Deaths for Births from January to December 2018. 2020.

27. OddD, WilliamsT, StoianovaS, RossouwG, FlemingP, LuytK. Newborn Health and Child Mortality Across England. JAMA Netw Open. 2023;6(10):e2338055.

28. Infection related deaths of children and young people in England. National Child Mortality Database (NCMD); 2023.

29. SimoncicV, DeguenS, EnauxC, VandentorrenS, Kihal-TalantikiteW. A Comprehensive Review on Social Inequalities and Pregnancy Outcome-Identification of Relevant Pathways and Mechanisms. Int J Environ Res Public Health. 2022;19(24).

30. BoghossianNS, GeraciM, LorchSA, PhibbsCS, EdwardsEM, HorbarJD. Racial and Ethnic Differences Over Time in Outcomes of Infants Born Less Than 30 Weeks’ Gestation. Pediatrics. 2019;144(3).

31. DagherRK, LinaresDE. A Critical Review on the Complex Interplay between Social Determinants of Health and Maternal and Infant Mortality. Children (Basel). 2022;9(3).

32. MichelM, AlbertiC, CarelJC, ChevreulK. Social inequalities in access to care at birth and neonatal mortality: an observational study. Arch Dis Child Fetal Neonatal Ed. 2022;107(4):380–5.

33. BaauwA, RosiekS, SlatteryB, ChinapawM, van HensbroekMB, van GoudoeverJB, et al.Pediatrician-experienced barriers in the medical care for refugee children in the Netherlands. Eur J Pediatr. 2018;177(7):995–1002.

34. KhanomA, AlanazyW, CouzensL, EvansBA, FaganL, FogartyR, et al.Asylum seekers’ and refugees’ experiences of accessing health care: a qualitative study. BJGP Open. 2021;5(6).

35. NicholsonE, McDonnellT, De BrúnA, BarrettM, BuryG, CollinsC, et al.Factors that influence family and parental preferences and decision making for unscheduled paediatric healthcare - systematic review. BMC Health Serv Res. 2020;20(1):663.

36. TohRKC, ShoreyS. Experiences and needs of women from ethnic minorities in maternity healthcare: A qualitative systematic review and meta-aggregation. Women Birth. 2023;36(1):30–8.

37. Inquiry into racial injustice in maternity care. Birthrights; 2022.

38. HorvatL, HoreyD, RomiosP, Kis-RigoJ. Cultural competence education for health professionals. Cochrane Database Syst Rev. 2014;2014(5):Cd009405.

39. RaziehC, PowellB, DrummondR, WardIL, MorganJ, GlickmanM, et al.Understanding the quality of ethnicity data recorded in health-related administrative data sources compared with Census 2021 in England. PLoS Med. 2025;22(2):e1004507.

