## Supplementary material for "ETHNICITY, DEPRIVATION, AND CHILDHOOD MORTALITY IN ENGLAND: A COHORT STUDY": eFigure 1

**eFigure 1. Unadjusted rate of death per 100,000 person years, and age at death (3 point smoothed using Gaussian weighting)**

Rate of death per 100,000 Children per Year (N.B. Base 2 log scale)

| Age at Death (Years) | White or White British | Asian or Asian British | Black or Black British | Mixed or Other |
| --- | --- | --- | --- | --- |
| <1 year | 2252.18 | 3885.86 | 4675.92 | 2341.94 |
| 1 | 617.326 | 1083.11 | 1232.63 | 690.485 |
| 2 | 136.497 | 264.03 | 233.872 | 180.191 |
| 3 | 98.4081 | 211.969 | 116.04 | 127.002 |
| 4 | 75.2012 | 160.984 | 126.918 | 98.6909 |
| 5 | 65.8815 | 139.491 | 127.711 | 106.738 |
| 6 | 61.6138 | 139.026 | 120.041 | 84.02 |
| 7 | 64.8864 | 106.511 | 70.8276 | 57.9988 |
| 8 | 63.5734 | 94.9295 | 85.0173 | 62.5985 |
| 9 | 58.9995 | 104.231 | 88.2948 | 75.0614 |
| 10 | 56.0262 | 88.8168 | 81.2055 | 77.3612 |
| 11 | 69.5762 | 112.15 | 98.3318 | 64.9563 |
| 12 | 83.202 | 134.055 | 124.396 | 88.9358 |
| 13 | 95.4439 | 158.397 | 147.687 | 122.554 |
| 14 | 110.717 | 164.078 | 164.959 | 141.179 |
| 15 | 142.158 | 176.654 | 192.418 | 146.921 |
| 16 | 169.948 | 207.396 | 234.719 | 153.91 |
| 17 | 229.304 | 233.972 | 242.033 | 214.257 |
