## Supplementary material for "ETHNICITY, DEPRIVATION, AND CHILDHOOD MORTALITY IN ENGLAND: A COHORT STUDY": eTable 1

**eTable 1. Characteristics of CYP deaths reported to NCMD in England between April 2019 and March 2023; split by ethnicity**

| **Measure** | **N** | **White** | **Asian** | **Black** | **Mixed** | **Other** | **p-value** |
| --- | --- | --- | --- | --- | --- | --- | --- |
| All Deaths | 12,142 | 7758 | 2242 | 1039 | 742 | 361 |  |
| Age (Categories) | 12,142 |  |  |  |  |  | <0.001 |
| 0-4 years | 8569 | 5374 (69.3%) | 1639 (73.1%) | 758 (73.0%) | 550 (74.1%) | 248 (68.7%) |  |
| 5-15 years | 2537 | 1643 (21.2%) | 463 (20.7%) | 202 (19.4%) | 141 (19.0%) | 88 (24.4%) |  |
| 16/17 years | 1036 | 741 (9.6%) | 140 (6.2%) | 79 (7.6%) | 51 (6.9%) | 25 (6.9%) |  |
| Age (Years) | 12,142 |  |  |  |  |  | <0.001 |
| <1 year | 7157 | 4506 (58.1%) | 1337 (59.6%) | 655 (63.0%) | 470 (63.3%) | 189 (52.4%) |  |
| 1 | 551 | 342 (4.4%) | 111 (5.0%) | 43 (4.1%) | 29 (3.9%) | 26 (7.2%) |  |
| 2 | 362 | 225 (2.9%) | 75 (3.4%) | 31 (3.0%) | 18 (2.4%) | 16 (4.4%) |  |
| 3 | 278 | 171 (2.2%) | 66 (2.9%) | 10 (1.0%) | 22 (3.0%) | 9 (2.5%) |  |
| 4 | 218 | 130 (1.7%) | 50 (2.2%) | 19 (1.8%) | 11 (1.5%) | 9 (2.2%) |  |
| 5 | 205 | 122 (1.6%) | 43 (1.9%) | 17 (1.6%) | 10 (1.4%) | 13 (3.6%) |  |
| 6 | 194 | 108 (1.4%) | 49 (2.2%) | 19 (1.8%) | 10 (1.4%) | 8 (2.2%) |  |
| 7 | 175 | 123 (1.6%) | 33 (1.5%) | 7 (0.7%) | 8 (1.1%) | NA |  |
| 8 | 184 | 123 (1.6%) | 30 (1.3%) | 14 (1.4%) | 13 (1.4%) | NA |  |
| 9 | 186 | 117 (1.5%) | 39 (1.7%) | 14 (1.4%) | 8 (1.1%) | 8 (2.2%) |  |
| 10 | 158 | 101 (1.3%) | 25 (1.1%) | 12 (1.25) | 13 (1.8%) | 7 (1.9%) |  |
| 11 | 207 | 137 (1.8%) | 39 (1.7%) | 16 (1.5%) | 12 (1.6%) | NA |  |
| 12 | 243 | 160 (2.1%) | 44 (2.0%) | 21 (2.0%) | 9 (1.2%) | 9 (2.5%) |  |
| 13 | 291 | 185 (2.4%) | 55 (2.5%) | 27 (2.4%) | 14 (1.9%) | 12 (3.2%) |  |
| 14 | 309 | 197 (2.5%) | 52 (2.3%) | 27 (2.6%) | 24 (3.2%) | 9 (2.5%) |  |
| 15 | 385 | 270 (3.7%) | 54 (2.4%) | 30 (2.9%) | 20 (2.7%) | 11 (3.0%) |  |
| 16 | 424 | 287 (3.7%) | 66 (2.9%) | 40 (3.9%) | 24 (3.2%) | 7 (1.9%) |  |
| 17 | 612 | 454 (5.9%) | 74 (3.3%) | 39 (3.8%) | 27 (3.6%) | 18 (5.0%) |  |
| Sex | 12,108 |  |  |  |  |  | 0.078 |
| Female | 5210 | 3277 (42.3%) | 1023 (45.7%) | 444 (42.9%) | 310 (42.2%) | 156 (43.2%) |  |
| Male | 6898 | 4463 (57.7%) | 1214 (54.3%) | 591 (57.1%) | 425 (57.8%) | 205 (56.8%) |  |
| Region | 12,142 |  |  |  |  |  | <0.001 |
| East Midlands | 1012 | 731 (9.4%) | 131 (5.8%) | 62 (6.0%) | 75 (10.1%) | 13 (3.6%) |  |
| East of England | 1165 | 837 (10.8%) | 144 (6.4%) | 78 (7.5%) | 77 (10.4%) | 29 (8.0%) |  |
| London | 2006 | 731 (9.4%) | 557 (24.8%) | 457 (44.0%) | 186 (25.1%) | 135 (37.1%) |  |
| North East | 602 | 499 (6.4%) | 58 (2.6%) | 21 (2.0%) | 8 (1.1%) | 167 (4.4%) |  |
| North West | 1762 | 1237 (15.9% | 335 (15.0%) | 80 (7.7%) | 68 (9.2%) | 42 (11.6%) |  |
| South East | 1640 | 1178 (15.2%) | 226 (10.1%) | 102 (9.8%) | 103 (13.9%) | 31 (8.6%) |  |
| South West | 914 | 792 (10.2%) | 31 (1.4%) | 21 (3.0%) | 42 (5.7%) | 18 (5.0%) |  |
| West Midlands | 1652 | 910 (11.7%) | 429 (19.1%) | 71 (13.2%) | 121 (16.3%) | 55 (15.2%) |  |
| Yorkshire and the Humber | 1329 | 843 (10.9%) | 331 (14.8%) | 71 (6.8%) | 62 (8.4%) | 22 (6.1%) |  |
| Deprivation Decile | 12,081 |  |  |  |  |  | <0.001 |
| 1 (Most Deprived) | 2315 | 1299 (16.8%) | 574 (25.6%) | 241 (23.3%) | 131 (17.7%) | 70 (20.4%) |  |
| 2 | 1817 | 979 (12.7%) | 423 (18.9%) | 230 (22.2%) | 126 (17.0%) | 59 (17.2%) |  |
| 3 | 1493 | 836 (10.8%) | 331 (14.8%) | 181 (17.5%) | 93 (12.6%) | 52 (15.2%) |  |
| 4 | 1256 | 757 (9.8%) | 250 (11.2%) | 130 (12.6%) | 79 (10.7%) | 40 (11.7%) |  |
| 5 | 1112 | 688 (8.9%) | 219 (9.8%) | 105 (10.1%) | 73 (9.9%) | 27 (7.9%) |  |
| 6 | 998 | 740 (9.6%) | 135 (6.0%) | 52 (5.0%) | 46 (6.2%) | 25 (7.3%) |  |
| 7 | 877 | 656 (8.5%) | 102 (4.6%) | 38 (3.7%) | 54 (7.3%) | 27 (7.9%) |  |
| 8 | 802 | 632 (8.2%) | 72 (3.2%) | 37 (3.6%) | 51 (6.9%) | 10 (2.9%) |  |
| 9 | 771 | 618 (8.0%) | 76 (3.4%) | 14 (1.4%) | 40 (5.4%) | 23 (6.7%) |  |
| 10 (Least Deprived) | 640 | 516 (6.7%) | 58 (2.6%) | 8 (0.8%) | 48 (6.5%) | 10 (2.9%) |  |
| Area | 12,085 |  |  |  |  |  | <0.001 |
| Urban | 10,705 | 6430 (83.3%) | 2219 (99.1%) | 1023 (98.7%) | 697 (94.1%) | 336 (98.0%) |  |
| Rural | 1380 | 1294 (15.8%) | 21 (0.9%) | 14 (1.4%) | 44 (5.9%) | 7 (2.0%) |  |

Values are number (%)

NA – Not available due to small numbers
