## Supplementary material for "ETHNICITY, DEPRIVATION, AND CHILDHOOD MORTALITY IN ENGLAND: A COHORT STUDY": eTable 2

**eTable 2. Number of deaths reported to NCMD in England between April 2019 and March 2023; split by ethnicity**

| **Measure** | **N** | **White** | **Asian** | **Black** | **Mixed** | **Other** | **p-value** |
| --- | --- | --- | --- | --- | --- | --- | --- |
| All Deaths | 12,142 | 7758 | 2242 | 1039 | 742 | 361 |  |
| Death by Cause | 11,872 |  |  |  |  |  | <0.001 |
| Malignancy | 999 | 717 (9.4%) | 125 (5.8%) | 69 (6.8%) | 49 (6.7%) | 39 (11.0%) |  |
| Preterm Birth | 2703 | 1691 (22.3%) | 477 (22.0%) | 287 (28.2%) | 182 (24.8%) | 39 (11.0%) |  |
| Intrapartum Event | 650 | 462 (6.1%) | 89 (4.1%) | 40 (3.9%) | 44 (6.0%) | 15 (4.2%) |  |
| Infection | 613 | 374 (4.9%) | 141 (6.5%) | 54 (5.3%) | 21 (2.9%) | 23 (6.5%) |  |
| Trauma | 773 | 519 (6.8%) | 96 (4.4%) | 82 (8.1%) | 54 (7.4%) | 22 (6.2%) |  |
| SUDIC | 3840 | 1261 (16.6%) | 209 (9.6%) | 147 (14.4%) | 139 (19.0%) | 37 (10.4%) |  |
| Underlying Disease | 1793 | 2180 (28.7%) | 1001 (46.1%) | 316 (31.0%) | 206 (28.1%) | 137 (38.6%) |  |
| Suicide | 451 | 350 (4.6%) | 32 (1.5%) | 22 (2.2%) | 33 (4.5%) | 14 (3.9%) |  |
| Substance Abuse | 50 | 42 (0.6%) | - | - | 5 (0.7%) | - |  |

Values are n(%)
