## Supplementary material for "ETHNICITY, DEPRIVATION, AND CHILDHOOD MORTALITY IN ENGLAND: A COHORT STUDY": eTable 3

**eTable 3. Incident Rate Ratio (IRR) of CYP death by ethnicity, stratified by age in years**

| **Measure** | **N** | **White** | **Asian** | **Black** | **Mixed** | **Other** | **p_interaction_** |
| --- | --- | --- | --- | --- | --- | --- | --- |
| Age (Years) | 12,142 |  |  |  |  |  | <0.001 |
| <1 year | 7157 | 1 (Ref) | 1.72 (1.62-1.83) | 2.08 (1.92-2.26) | 0.89 (0.81-0.98) | 1.17 (1.01-1.36) |  |
| 1 | 551 | 1 (Ref) | 1.83 (1.48-2.27) | 1.72 (1.25-2.36) | 0.73 (0.50-1.07) | 2.03 (1.36-3.02) |  |
| 2 | 362 | 1 (Ref) | 1.92 (1.48-2.49) | 1.88 (1.29-2.74) | 0.71 (0.44-1.14) | 1.89 (1.14-3.14) |  |
| 3 | 278 | 1 (Ref) | 2.24 (1.68-2.97) | 0.80 (0.42-1.52) | 1.18 (0.76-1.84) | 1.43 (0.73-2.79) |  |
| 4 | 218 | 1 (Ref) | 2.14 (1.54-2.96) | 1.95 (1.20-3.15) | 0.79 (0.42-1.45) | 1.63 (0.80-3.33) |  |
| 5 | 205 | 1 (Ref) | 2.02 (1.43-2.86) | 1.85 (1.11-3.07) | 0.79 (0.41-1.50) | 2.77 (1.56-4.90) |  |
| 6 | 194 | 1 (Ref) | 2.53 (1.81-3.55) | 2.32 (1.42-3.77) | 0.91 (0.47-1.73) | 1.93 (0.94-3.96) |  |
| 7 | 175 | 1 (Ref) | 1.50 (1.02-2.21) | 0.73 (0.34-1.57) | 0.66 (0.32-1.35) | 0.85 (0.31-2.31) |  |
| 8 | 184 | 1 (Ref) | 1.39 (0.93-2.07) | 1.45 (0.84-2.53) | 1.08 (0.61-1.92) | 0.87 (0.32-2.35) |  |
| 9 | 186 | 1 (Ref) | 1.93 (1.34-2.78) | 1.52 (0.87-2.64) | 0.73 (0.35-1.49) | 1.83 (0.89-3.74) |  |
| 10 | 158 | 1 (Ref) | 1.46 (0.94-2.26) | 1.45 (0.80-2.64) | 1.40 (0.79-2.50) | 1.84 (0.85-3.95) |  |
| 11 | 207 | 1 (Ref) | 1.65 (1.16-2.36) | 1.38 (0.82-2.32) | 0.97 (0.54-1.76) | 0.57 (0.18-1.79) |  |
| 12 | 243 | 1 (Ref) | 1.57 (1.13-2.20) | 1.50 (0.95-2.37) | 0.63 (0.32-1.24) | 1.49 (0.76-2.91) |  |
| 13 | 291 | 1 (Ref) | 1.71 (1.27-2.31) | 1.55 (1.02-2.36) | 0.87 (0.51-1.50) | 1.75 (0.98-3.14) |  |
| 14 | 309 | 1 (Ref) | 1.54 (1.14-2.09) | 1.56 (1.04-2.33) | 1.46 (0.96-2.23 | 1.23 (0.64-2.44) |  |
| 15 | 385 | 1 (Ref) | 1.17 (0.87-1.56) | 1.23 (0.86-1.84) | 0.93 (0.59-1.47) | 1.11 (0.61-2.03) |  |
| 16 | 424 | 1 (Ref) | 1.33 (1.02-1.74) | 1.57 (1.13-2.19) | 1.07 (0.71-1.62) | 0.65 (0.31-1.38) |  |
| 17 | 612 | 1 (Ref) | 0.97 (0.76-1.24) | 0.97 (0.70-1.35) | 0.81 (0.55-1.20) | 1.08 (0.67-1.72) |  |

Values are IRR (95% CI)

NA – Not available due to small numbers
