## Supplementary material for "ETHNICITY, DEPRIVATION, AND CHILDHOOD MORTALITY IN ENGLAND: A COHORT STUDY": eTable 4

**eTable 4. Number of deaths reported to NCMD in England between April 2019 and March 2023; split by detailed measures of ethnicity (n=12,122)**

| **Measure** | **N** | White | | | | Asian or Asian British | | | | | Black or Black British | | | Mixed | | | | Other | | p-value |
| --- | --- | --- | --- | --- | --- | --- | --- | --- | --- | --- | --- | --- | --- | --- | --- | --- | --- | --- | --- | --- |
|  | **N** | British | Irish | Gypsy or Irish traveller | Other (Inc Roma, Gypsy or Irish traveller) | Bangladeshi | Chinese | Indian | Pakistani | Other | African | Caribbean | Other | White and Asian | White and Black African | White and Black Caribbean | Other | Arab | Other |  |
| All Deaths |  | 6781 | 68 | 31 | 871 | 262 | 40 | 405 | 1113 | 417 | 729 | 156 | 154 | 180 | 103 | 225 | 234 | 59 | 294 |  |
| Death by Cause |  |  |  |  |  |  |  |  |  |  |  |  |  |  |  |  |  |  |  | <0.001 |
| Malignancy |  | 639 (9.6%) | NA | NA | 72 (8.4%) | 6 (2.3%) | NA | 111 (27.9%) | 204 (19.2%) | 87 (21.4%) | 196 (27.6%) | 10 (6.4%) | 9 (5.9%) | NA | NA | NA | NA | NA | 30 (10.3%) |  |
| Preterm Birth |  | 1434 (21.6%) | NA | NA | 238 (27.8%) | 69 (26.7%) | NA | 111 (27.9%) | 204 (19.2%) | 87 (21.4%) | 196 (27.6%) | 55 (35.3%) | 36 (23.5%) | NA | NA | NA | NA | NA | 61 (21.0%) |  |
| Intrapartum Event |  | 390 (5.9%) | NA | NA | 69 (8.1%) | 11 (4.3%) | NA | 28 (7.0%) | 34 (3.2%) | 16 (3.9%) | 27 (3.8%) | 9 (5.8%) | NA | NA | NA | NA | NA | NA | 13 (4.5%) |  |
| Infection |  | 330 (5.0%) | NA | NA | 42 (4.9%) | 16 (6.2%) | NA | 33 (8.3%) | 67 (6.3%) | 24 (5.9%) | 37 (5.2%) | 10 (6.4%) | 8 (5.2%) | NA | NA | NA | NA | NA | 17 (5.9%) |  |
| Trauma |  | 446 (6.7%) | NA | NA | 56 (6.6%) | 8 (3.1%) | NA | NA | 21 (5.3%) | 42 (3.9%) | 21 (5.2%) | 17 (10.9%) | 18 (11.8%) | NA | NA | NA | NA | NA | 20 (6.9%) |  |
| SUDIC |  | 1157 (17.4%) | NA | NA | 89 (10.4%) | 27 (10.5%) | NA | 45 (11.3%) | 85 (8.0%) | 44 (10.8%) | 104 (14.6% | 17 (10.9%) | 27 (17.7%) | NA | NA | NA | NA | NA | 27 (9.3%) |  |
| Underlying Disease |  | 1890 (28.5%) | NA | NA | 255 (29.8%) | 117 (45.4%) | NA | 128 (32.2%) | 566 (53.2%) | 174 (42.8%) | 232 (32.6%) | 36 (23.1%) | 48 (31.4%) | NA | NA | NA | NA | NA | 107 (36.9%) |  |
| Suicide |  | NA | NA | NA | NA | NA | NA | NA | NA | NA | NA | NA | NA | NA | NA | NA | NA | NA | NA |  |
| Substance Abuse |  | NA | NA | NA | NA | NA | NA | NA | NA | NA | NA | NA | NA | NA | NA | NA | NA | NA | NA |  |

Values are n(%)
